# Clinically Applicable Deep Learning Strategy for Pulmonary Nodule Risk Prediction: Insights into HONORS

**DOI:** 10.1101/2020.02.03.20020297

**Authors:** Wenhui Lv, Yang Wang, Changsheng Zhou, Sheng Huang, Xiangming Fang, Qiuzhen Xu, Qirui Zhang, Chuxi Huang, Xinyu Li, Zhen Zhou, Yizhou Yu, Yizhou Wang, Mengjie Lu, Qiang Xu, Xiuli Li, Haoliang Lin, Xiaofan Lu, Qinmei Xu, Jing Sun, Yuxia Tang, Yong Song, Fangrong Yan, Bing Zhang, Zhen Cheng, Longjiang Zhang, Guangming Lu

## Abstract

**Background and Purpose:** Limited optimization was clinically applicable for reducing missed diagnosis, misdiagnosis and inter-reader variability in pulmonary nodule diagnosis. We aimed to propose a deep learning-based algorithm and a practical strategy to better stratify the risk of pulmonary nodules, thus reducing medical errors and optimizing the clinical workflow.

**Materials and Methods:** A total of 2,348 pulmonary nodules (1,215 with lung cancer) containing screened nodules from National Lung Cancer Screening Trial (NLST) and incidentally detected nodules from Jinling Hospital (JLH) were used to train and evaluate a deep learning algorithm, Filter-guided pyramid network (FGP-NET). Internal and external test of FGP-NET were performed on two independent datasets (n=542). The performance of FGP-NET at Youden point which maximizing the Youden index was compared with 126 board-certificated radiologists. We further proposed Hierarchical Ordered Network ORiented Strategy (HONORS), which manipulates the emphasis either on sensitivity or specificity to target risk-stratified clinical scenarios, directly making decisions for some patients.

**Results:** FGP-NET achieved a high area under the curve (AUC) of 0.969 and 0.855 for internal and external testing, and was comparable or even outperformed the radiologists when considering sensitivity. HONORS-guided FGP-NET identified benign nodules with a high sensitivity (95.5%) in the screening scenario, and demonstrated satisfactory performance for the rest ambiguous nodules with 0.945 of AUC by the Youden point. FGP-NET also detected lung cancer with a high specificity of 94.5% in routine diagnostic scenario; an AUC of 0.809 was achieved for the rest nodules.

**Conclusion:** The combination of HONORS and FGP-NET provides well-organized stratification for pulmonary nodules and also offers the potential for reducing medical errors.

**Highlights:**

- Pulmonary nodules were managed for both screening and diagnostic scenarios
- Proposal of a hierarchical strategy for targeting risk-stratified clinical scenarios
- A large scale Human-deep learning contest for reliable performance evaluation

## Introduction

Lung cancer is one of the commonest cancers and the leading cause of cancer mortality worldwide ^1,2^. Survival rate is hugely influenced by stage at diagnosis and early diagnosis is full of challenges ^3^. Every step from screening to final diagnosis is indispensable which makes the entire process time- and labor-consuming. For better management and diagnosis of pulmonary nodules, Lung-RADs guidelines ^4^, recommendations from Fleschnier society ^5^ have been proposed. However, the lack of awareness of these guidelines and the inter-reader variability have limited the broader use of them ^6,7^. Besides, the detection of pulmonary nodules is rising up dramatically and physicians are experiencing burnout at increasing rate. Excessive workload is resulting in diagnostic errors including missed diagnosis, and misdiagnosis ^8^. Therefore, there is an unmet need for relieving working pressure of physicians and reducing the incidence of such medical errors. To the best of our knowledge, limited literature has delved into this place for a clinically applicable optimization.

The desire to improve the efficacy and efficiency of clinical care continues to drive multiple innovations into practice, including deep learning (DL). In recent, DL has infiltrated the optimization and streamlining of clinical workflows, quietly improving, changing and reconstructing the way health care works ^9-13^, especially for pulmonary nodules management ^12,14^. However, most of previous works focused on the screening population, and studies based on incidentally detected nodules in routine diagnostic scenario with higher risk were limited.

In this study, we reasoned that DL would play distinct roles in the risk-stratified clinical scenarios for pulmonary nodule screening and diagnosis. Therefore, we proposed a clinically applicable DL-based algorithm—Filter-guided pyramid network (FGP-NET), and a practical strategy—Hierarchical-Ordered Network-ORiented Strategy (HONORS), which involves two steps for two different clinical scenarios (*i.e*., screening and routine diagnostic scenarios) (Figure 1a). The benign nodules or lung cancer can be accurately identified in screening and routine diagnostic scenarios in step-1 and further stratification of ambiguous nodules was performed to aid clinical decision making in step-2. Consequently, HONORS would directly make decision for some patients without any human intervention and assist physicians to better manage other ambiguous nodules. It has great potential to provide well-organized management for pulmonary nodules by optimizing the clinical workflow and reduce medical errors through rapid and accurate image interpretation.

**Figure 1:**
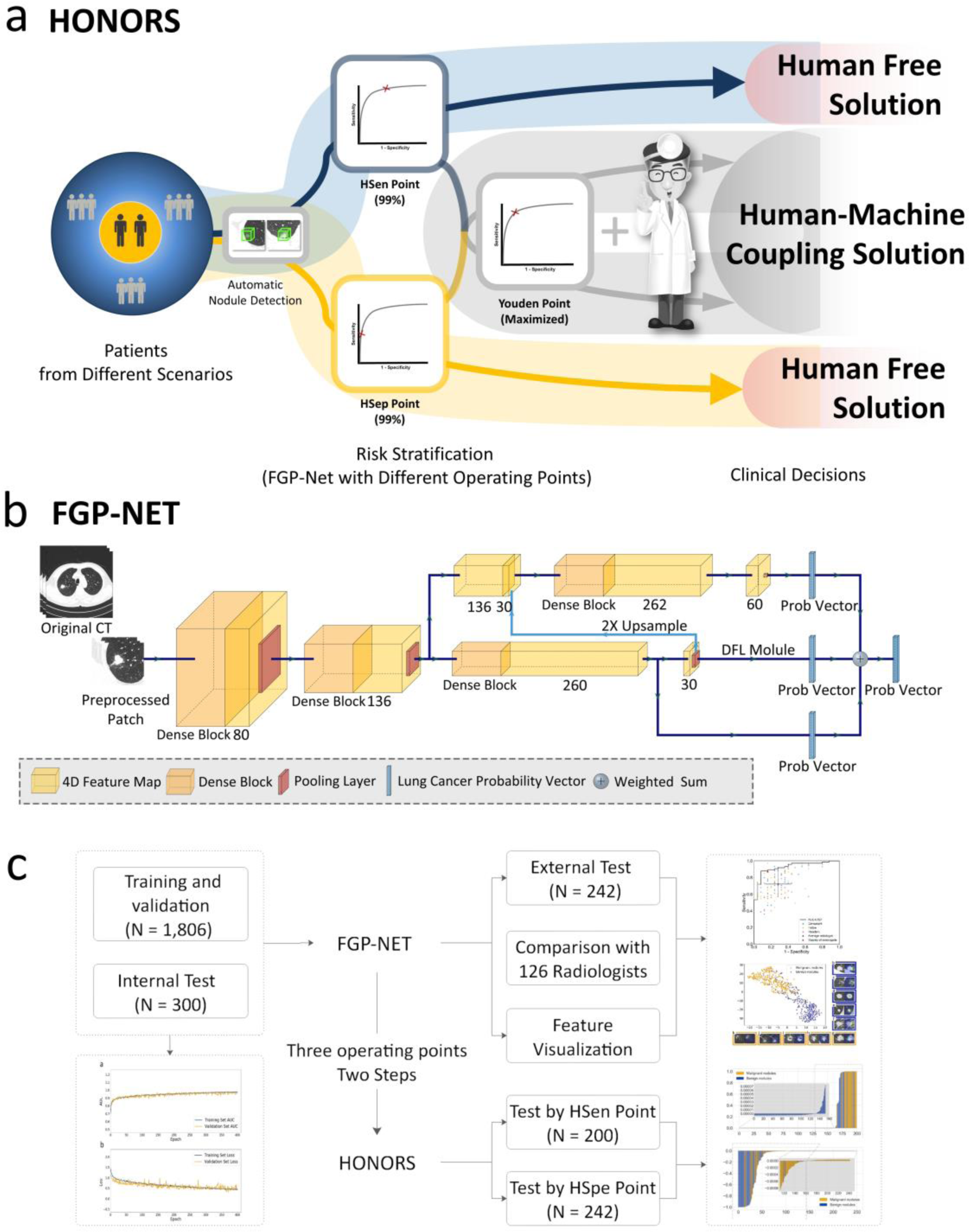
The framework of the two-step hierarchical strategy and the overall flow chart of this study. a) We proposed HONORS, a hierarchical strategy for pulmonary nodules risk stratification in different clinical scenarios, involves three operating points of FGP-NET, as well as a two-step method. The first step includes the blue or yellow translucent paths that focus on either sensitivity or specificity for screening or routine diagnosis scenarios, resulting in a portion of nodules of interest with high precision being detected without any intervention of physicians (radiologists) — “Human Free”. Further, under a Youden point in the second step (maximizing the Youden index), the remaining ‘ambiguous nodules’ are differentiated into benign and malignant ones by FGP-NET but require final confirmation by physicians (radiologists), which indicated a “Human-Machine Coupling Solution” mode. b) Schematic diagram of FGP-NET. c) The overall flow chart of this study, including the different clinical scenarios and the corresponding sample size.

## Methods

### Datasets

Ethical approval was obtained for this retrospective study, and informed consent was waived for reviewing patients’ medical records.

For training and validation of FGP-NET, we incorporated both incidentally detected and screened nodules from Jinling Hospital (JLH) and National Lung Cancer Screening Trial (NLST), respectively. We retrospectively analyzed 16,801 patients who underwent surgery or biopsy due to lung lesions from May 2009 to June 2018 in Jinling hospital. Nodules were classified as either malignant or benign according to the World Health Organization (WHO) classification of lung tumors in 2015 ^15^. Finally, a total of 1,286 nodules (935 malignant nodules and 351 benign nodules) found in 1,238 patients (591 males and 647 females; 56.48 ± 11.50 years of age [mean ± standard deviation (SD)]) were included. We analyzed 1,060 patients confirmed as having lung cancer and 5,275 patients as not in the NLST. A total of 820 nodules (93 malignant nodules and 727 benign nodules) found in 442 participants (244 males and 198 females; 61.71 ± 5.32 years of age) were included. All participants enrolled in NLST signed an informed consent developed and approved by the screening centers’ Institutional Review Board (IRB), the National Cancer Institute (NCI) IRB and the Westat IRB. The data and images were obtained from the National Cancer Institute Cancer Data Access System ^16^.

We also conducted an external test of FGP-NET using a multi-center set. Multi-center set contained consecutive 235 patients (104 males and 131 females; 58.48 ± 10.75 years of age) with 242 incidentally detected nodules (187 malignant nodules and 55 benign nodules) from three tertiary hospitals. These patients all underwent biopsy or surgery. Details for the data curation are available in the Supplementary Information, Inclusion and exclusion criteria. All patients included in this study underwent nonenhanced chest computed tomography (CT) and the CT images were reconstructed with a thickness less than 2.5 mm (see more details of acquisition and reconstruction parameters in Supplementary Table 1).

### Annotation and preprocessing

Automatic nodule detection was performed using the Dr. wise platform ^17^ (the detection network could be found in the supplementary information, Pulmonary nodule detection network) and the geometric centers of the enrolled nodules were further revised by two radiologists. Due to the differences in pixel spacing and slice thickness, the CT images were subsequently linearly interpolated into 3D isotropic images with voxel spacing of 0.6 × 0.6 × 0.6 mm3. Since the datasets were relatively small compared to traditional image classification datasets for deep learning such as cifar10^18^ and ImageNet^19^, we conducted heavy data augmentations on all initially generated image patches (a size of 128 × 128 × 128) containing the nodules, e.g., 0-360 degree of random rotation, random zooming in or zooming out, random cropping and flipping. Processed patches were cropped to a size of 48 × 48 × 48 and used for algorithm training.

### Development and training of FGP-NET

JLH and NLST dataset were randomly assigned into one of the following three sets: training set (JLH, 1086 nodules; NLST, 520 nodules) for optimizing network weights, validation set (JLH, 100 nodules; NLST, 100 nodules) for deciding the values of hyperparameters and internal test set (JLH, 100 nodules; NLST, 200 nodules) for evaluating the performance. The patients in the training, validation and test sets were exclusive to each of the other data sets. Pulmonary nodule evaluation on chest CT is of great challenge partly due to the significant overlap and complicated interaction among visual features. To this end, we aimed to capture both local and global features and their interactive relationships to better represent the nodule. We designed a new U-shape network structure named FGP-Net which could produce a feature pyramid for different-sized local features (Figure 1b). By concatenating the clear attention map distilled by local feature extractors and raw feature maps from early stage of network, FGP-Net was able to keep high resolution details to describe small-sized local features and to use the accurate localization of large ones to guide small ones. Additionally, we added one more backbone module to dig complex semantic information and relationships between these various local ones. We applied Densely Connected Convolutional Network (DenseNet) ^20^ as our backbone network and Discriminative Filter Learning (DFL) ^21^ modules as local feature extractors for details of nodules. DenseNet was one of the state-of-art network structures in computer vision. The core of Densenet was a so-called dense block, in which every layer was connected directly to all the subsequent layers. DFL Module was a two-stream structure, enabling network to detect discriminative mid-level patches under weakly supervised condition.

FGP-Net was trained by using a batch size of 4 for 400 epochs and learning rate of 0.0001, with adam optimizer and 0.0001 weight decay. After 400 epochs, the training process was stopped due to the convergence of both the area under the curve (AUC) and cross-entropy loss (Supplementary Figure 1).

### Validation of FGP-NET

FGP-Net generated continuous numbers between 0 and 1 for nodule risk stratification, being consistent with the malignancy probability of the nodules, named ‘malignancy score’. Receiver operating characteristic (ROC) curves were generated by plotting sensitivity and 1 - specificity, and AUCs were calculated to evaluate the discrimination ^22^. Three operating points were chosen for FGP-NET on the validation set; a high sensitivity (HSen)/specificity (HSpe) point corresponded to a sensitivity/specificity of 99% to stratify the benign/malignant nodules with high precision and a Youden point maximized the value of the Youden index for generic discrimination between malignant and benign nodules.

### Human-DL contest between FGP-NET and 126 radiologists

We recruited 126 radiologists from 41 hospitals in Jiangsu Province, P.R. China, to rate the nodules on the JLH test set. Among them, 41 (32.5%) radiologists are ‘resident’, 42 (33.3%) are ‘fellow’, and 43 (34.1%) are ‘consultant’. The rating task was held as a competition during an academic conference in China in 2018. All radiologists participating in this competition were informed of and agreed with the rationale and goals of this competition and participated on a voluntary basis. They individually reviewed the nodules using an image viewer with capabilities for picture archiving and communication system features (such as zoom, window leveling, and contrast adjustment) and rated the nodules from 1 to 4 (4 denotes the highest malignancy probability). The corresponding sets were defined as S1, S2, S3 and S4, respectively. To keep up with the input of FGP-NET, radiologists were only informed with the precise location of the nodules while blinded to the medical history and pathological results.

The performance of radiologists was evaluated at the average level and majority level (voting the scores rated by 126 radiologists) when compared with FGP-NET. S3, S4 were defined as malignant and S1, S2 were defined as benign. Sensitivity and specificity were evaluated and the 95% confidence intervals (CIs) of them were calculated by Wilson score CIs ^23^. Analysis of Variance (ANOVA) test was used to evaluate the performance of radiologists with different experience. Cohen’s κ coefficient (κw) was used to evaluate inter-observer agreement. The interobserver agreement between radiologists’ majority opinion and FGP-NET, radiologists’ majority opinion and each radiologist was examined. The interobserver agreement of radiologists in three groups with different experience was also examined. We categorized κ coefficients as follows: poor (0 < κw ≤ 0.20), fair (0.20 < κw ≤ 0.40), moderate (0.40 < κw ≤ 0.60), good (0.60 < κw ≤ 0.80), and excellent (0.80 < κw ≤ 1.00). To evaluate the impact of rating time on the performance, Spearman’s correlation coefficients between the rating time and sensitivity, specificity of radiologists were calculated. All statistical tests used in this study were 2-sided and a P-value less than .05 was considered significant. The statistics analyses were carried out using R software (version 3.5.3; http://www.Rproject.org).

### Interpretation of learned features

Given the black box property of DL, we further conducted a two-way feature interpretation to explore whether FGP-NET learned solid and effective features. More specifically, T-distributed Stochastic Neighborhood Embedding (t-SNE) ^24^ and probability heat-map were applied for visualization of global features and local features (Supplementary Information, Feature visualization of FGP-Net).

### Evaluation of HONORS

To validate the performance of HONORS in screening and routine diagnostic scenarios, we simulated the its application on NLST test set and multi-center set. In the first step, FGP-Net at HSen or HSpe point stratified the benign nodules or lung cancer in the NLST test set and multi-center set, respectively. The precision of the stratified nodules was evaluated using negative predictive value (NPV) and positive predictive value (PPV), respectively. In the second step, FGPNet at the Youden point stratified the rest ambiguous nodules and sensitivity and specificity were evaluated at this step. We further compared the performance between HONORS and 126 radiologists in the incidentally detected nodules using JLH test set (Supplementary information, Comparison of HONORS with radiologists). In addition to the two-step way targeted on different scenarios, HONORS can also be realized in a three-step way regardless of scenarios (Supplementary Figure 2). In the first step, FPG-NET at HSen point stratified the benign nodules; in the second step, FPG-NET at HSpe point stratified the lung cancer; and in the third step, FPG-NET at Youden point stratified the rest ambiguous nodules.

## Results

### Overview of the study design and results

A total of 2,348 pulmonary nodules (1,215 malignant nodules) containing screened and incidentally detected nodules found by chest CT were used to train and evaluate our DL algorithm, FGP-NET (the network structure of it could be found in Figure 1b). Detailed pathological types of different datasets are summarized in Supplementary Table 2. The entire workflow of this study is delineated in Figure 1c, including: i) the development and performance evaluation of FGP-NET, ii) a large scale Human-DL contest, iii) feature visualization, iv) proposal and validation of a novel two-step hierarchical strategy named HONORS for clinical application of FGP-NET in different scenarios.

### The construction and performance evaluation of FGP-NET

To assess the general performance of FPG-NET on the risk stratification of pulmonary nodules, we selected the optimal operating point, Youden point, which maximized the Youden index to weight both sensitivity and specificity (grey path in Figure 1a). FPG-NET achieved a dramatically high AUC of 0.969 (95% CI: 0.943-0.986) in the internal test set with a sensitivity of 93.8% (95% CI: 86.8-97.1%) and a specificity of 89.2% (95% CI: 84.0-92.8%) at the Youden point (Table 1). When considering the external test set, an AUC of 0.855 (95% CI: 0.804-0.896) and a high sensitivity of 96.3% (95% CI, 92.3-98.2%) were also achieved but the specificity decreased to 47.3% (95% CI: 34.1-60.2%), which may arise from the different nodule distribution, scanning and reconstruction parameters (Supplementary Table 1, 3).

**Table 1:** Performance of FGP-NET during internal and external test.

| Dataset | Prevalence (%) | AUC<sup>a</sup> | Sens (%)<sup>b</sup> | Spec (%)<sup>c</sup> | PPV(%)<sup>d</sup> | NPV(%)<sup>e</sup> |
| --- | --- | --- | --- | --- | --- | --- |
| Internal test <sup>f</sup> | 32.3 | 0.969 | 93.8 | 89.2 | 80.5 | 96.8 |
| JLH | 75.0 | 0.927 | 93.3 | 64.0 | 88.6 | 76.2 |
| NLST | 11.0 | 0.967 | 95.5 | 92.7 | 61.8 | 99.4 |
| External test <sup>g</sup> | 77.3 | 0.855 | 96.3 | 47.3 | 86.1 | 78.8 |

<sup>a</sup> AUC, area under the curve;
<sup>b</sup> Sens, sensitivity;
<sup>c</sup> Spec, specificity;
<sup>d</sup> PPV, positive predictive value;
<sup>e</sup> NPV, negative predictive value;
<sup>f</sup> Internal test set includes JLH (Jinling Hospital) dataset and NLST (National Lung Cancer Screening Trial) dataset;
<sup>g</sup> External test includes a multi-center test set.

### Human-DL contest between FGP-NET and 126 radiologists

We further investigated whether the FGP-NET was comparable or even superior to radiologists. To this end, 126 radiologists were recruited to participate in a Human-DL contest using JLH test set (see more details in METHODS). When assessing the decisions of each radiologist independently, the average sensitivity and specificity were 72.2 ± 15.1% and 71.7 ± 15.5%, respectively; while dramatically higher sensitivity (93.3%, 95% CI: 84.8-97.1%) but a relatively inferior specificity (64.0%, 95% CI: 43.3-79.8%) were achieved by FGP-NET (Figure 2a). Besides, FGP-NET had an advantage in sensitivity even compared with the consultant group (Supplementary Table 4). We then integrated the decisions of 126 radiologists through voting, and the sensitivity, specificity of radiologists’ majority opinion was computed to be 86.7% (95% CI: 76.8-92.6%) and 84.0% (95% CI: 64.1-93.6%), respectively (Figure 2a). Both of FGP-NET and radiologists’ majority opinion misdiagnosed 14 out of 100 nodules. These results indicated that the proposed FGP-NET was comparable or even outperformed the human radiologists when considering sensitivity.

**Figure 2:**
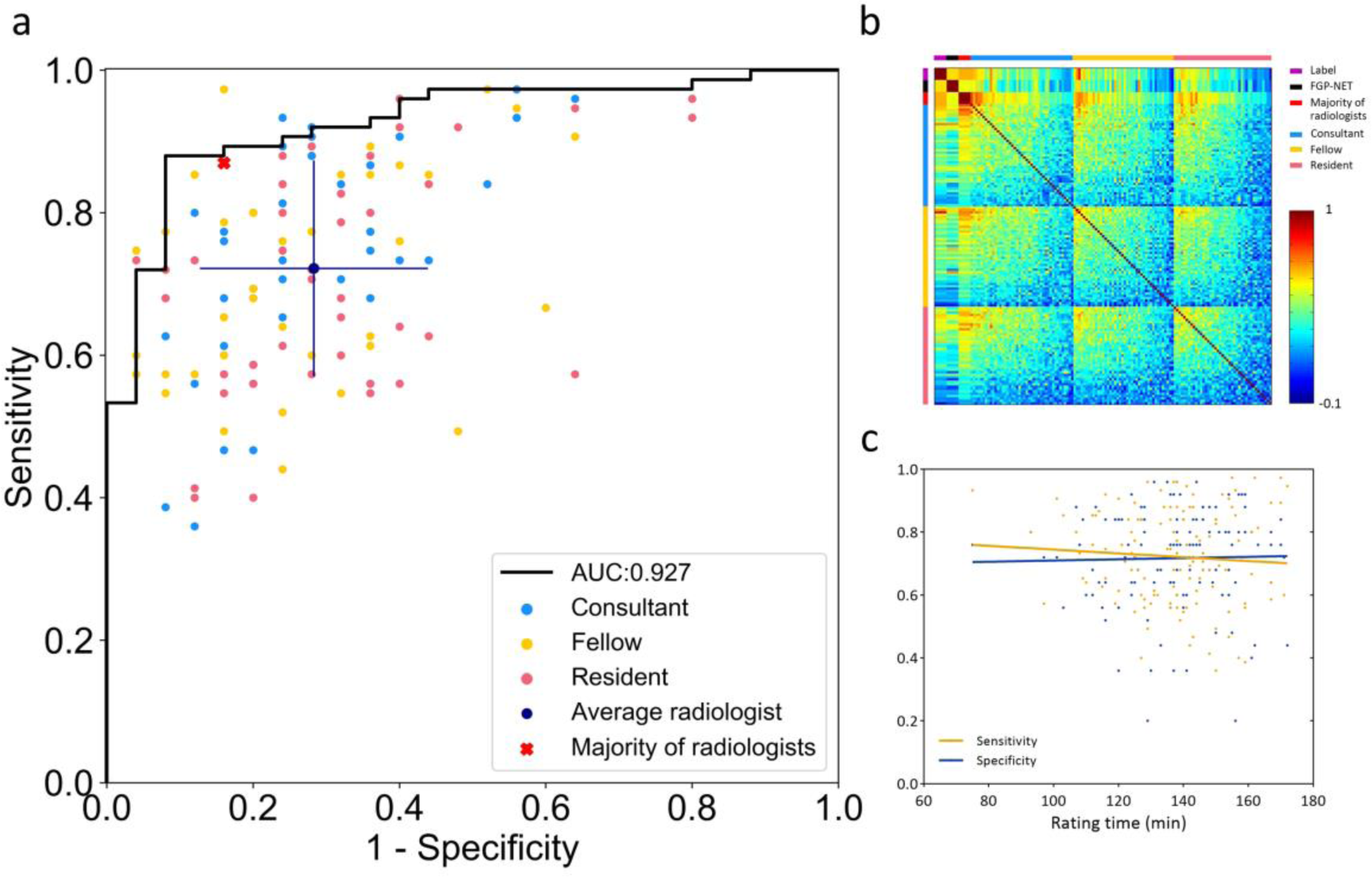
Performance comparison between FGP-NET and radiologists. a) We conducted a comparison study with a group of 126 radiologists. They reviewed and rated the nodules from 1 to 4 (4 denotes the highest malignancy). Performances of FGP-NET (black line) versus all radiologists (lake blue, yellow and pink points) are presented. Each point represents a combination of sensitivity and specificity for a radiologist. The indigo blue point represents the average performance of the radiologists, with error bars denoting the SD. The red cross mark represents the sensitivity and specificity of radiologist’ majority opinion (voting the score rated by 126 radiologists). b) The inter-observer agreement among label, FGP-NET, majority of radiologists and each radiologist. Inter-observer agreement between 126 radiologists was fair (κw = 0.315 ± 0.139). Although the inter-observer agreement grew with increased experience, inter-observer variability was huge even in consultants in pulmonary nodule diagnosis. c) Correlation analyses between the sensitivity, specificity and the rating time of each radiologist are shown.

Inter-observer agreement among 126 radiologists was fair (κw = 0.315 ± 0.139), and the average κw in resident, fellow and consultant groups were 0.278 ± 0.140, 0.315 ± 0.136 and 0.349 ± 0.131, respectively (Figure 2b). Although the inter-observer agreement grew with increased experience, it was still at a relatively low level even in consultant group (κw < 0.4). We found that the inter-observer agreement between the radiologists’ majority opinion and FGP-NET (κw = 0.538) was moderate, which outperformed 78 out of 126 (61.9%) radiologists. A total of 18 nodules was misdiagnosed by radiologists’ majority opinion and FGP-NET inconsistently, accounting for 9 each (Supplementary Figure 3). Interestingly, most misdiagnosed nodules of FGP-NET were granuloma which manifested as solid nodules on CT; while radiologists tended to misdiagnose adenocarcinoma that manifested as sub-solid nodules. The amount of time consumed by radiologists during the contest was not relevant with the ultimate sensitivity and specificity (all, *P* > 0.05; Figure 2c).

### Interpretation of Features Learned by FGP-NET

DL is frequently referred to as a black box—data goes in, decisions come out, but the processes between input and output are opaque ^25^. It is crucial to enable the black box to be opened, and thus, we interpreted the global features by using t-SNE which is particularly well suited for the visualization of high-dimensional data. The t-SNE demonstrated clear separation among the malignant and benign nodules (Figure 3a). We further visualized the local features via probability heat-map. To be specific, these attribution regions were mainly located in benign features for those benign nodules. The regions indicated that FGP-NET may focus on the center of the nodule or satellite lesions outside the nodule (Figure 3b-3f). Nevertheless, to malignant nodules, regions were predominately situated in malignant features, suggesting that FGP-NET may concentrate on the irregular margin of the nodule and solid component within part-solid nodules (Figure 3g-3k). These attribution regions were certainly consistent with the visual observation by radiologists.

**Figure 3:**
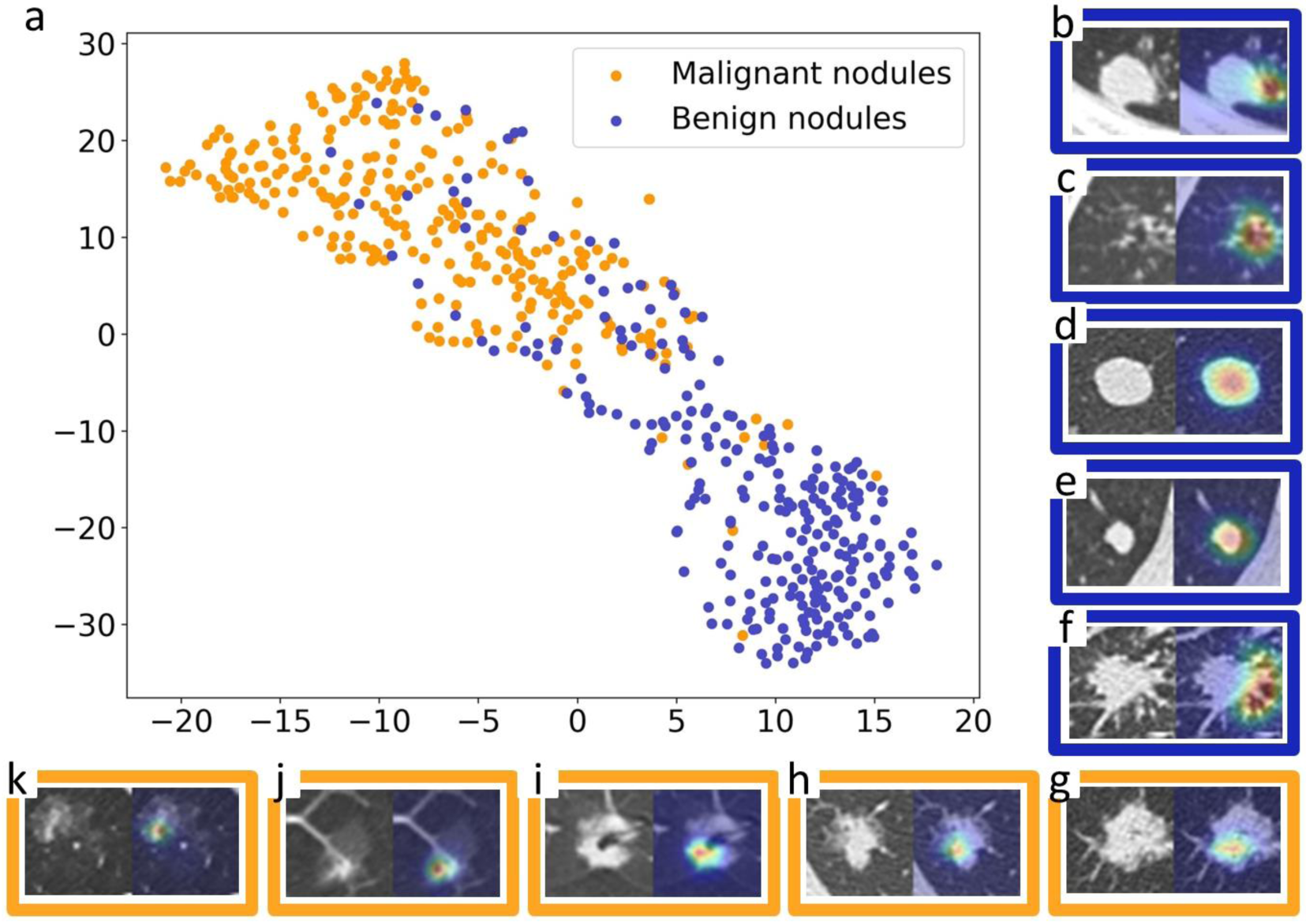
Opening the black box of FGP-NET by both t-SNE and probability heat-maps. a) t-SNE visualization of features extracted from the last fully-connected layer of the global branch of FGP-NET. Global image features learned by FGP-NET were visualized as a 2-dimensional scatter plot, with each point corresponding to a nodule that derives from either JLH, NLST or Multi-center dataset. Heat-maps of sample images reveal the regions that most positively influence the prediction of FGP-NET. The left column represents the central axial slice of the 48 × 48 patch network input. The right column represents the overlaid activation region, with red and blue showing the highest and lowest contributions, respectively. In benign nodules, activated maps were mainly centrally located in benign features (b-f). b-c) Granuloma, the activated region was in the middle of the satellite lesions. d-e) Hamartoma, the activated region was in the center of nodule. f) Inflammatory pseudotumor, the activated region was long speculation. In malignant nodules, they were mainly off-center located in malignant features (g-k).

### Proposal of HONORS and performance evaluation in different clinical scenarios

#### Proposal of HONORS

Although FGP-NET outperformed the radiologists in risk stratification of pulmonary nodules, this does not eliminate the role of radiologists in making the final decision. Therefore, we further proposed HONORS, a novel two-step hierarchical strategy for clinical application of FGP-NET. HONORS included a high sensitivity (Hsen) and high specificity (HSpe) point based on FGP-NET which manipulated the emphasis on either high sensitivity or specificity in screened and incidentally detected nodules, respectively. It enabled FGP-NET to make direct decisions for a portion of patients. In addition, Youden point was chosen to assist the diagnosis of rest ambiguous nodules. HONORS was applied to NLST test set and multi-center set to appraise its performance. Additionally, a three-step way to realize HONORS was also evaluated in this context (Supplementary Figure 2 and Supplementary Table 5, 6). We finally chose the two-step way as our main solution due to the good operability and specific target on different clinical scenarios.

#### Evaluation of HONORS in screening scenario

To screened nodules in screening scenario (blue path in Figure 1a), we deployed FGP-NET at a HSen point on NLST test set (n = 200) which represented screened population. A high sensitivity of 95.5% (95% CI: 76.8-99.2%) and a specificity of 72.5% (95% CI: 65.3-78.5%) were achieved in step-1 where FGP-NET stratified 130 nodules as benign (Figure 4a, c). In step-2, FGP-NET at the Youden point achieved a sensitivity of 100.0% (95% CI: 93.1-100.0%) and a specificity of 73.5% (95% CI: 59.1-83.8%) for the further stratification of ambiguous nodules (Figure 4b, c).

**Figure 4:**
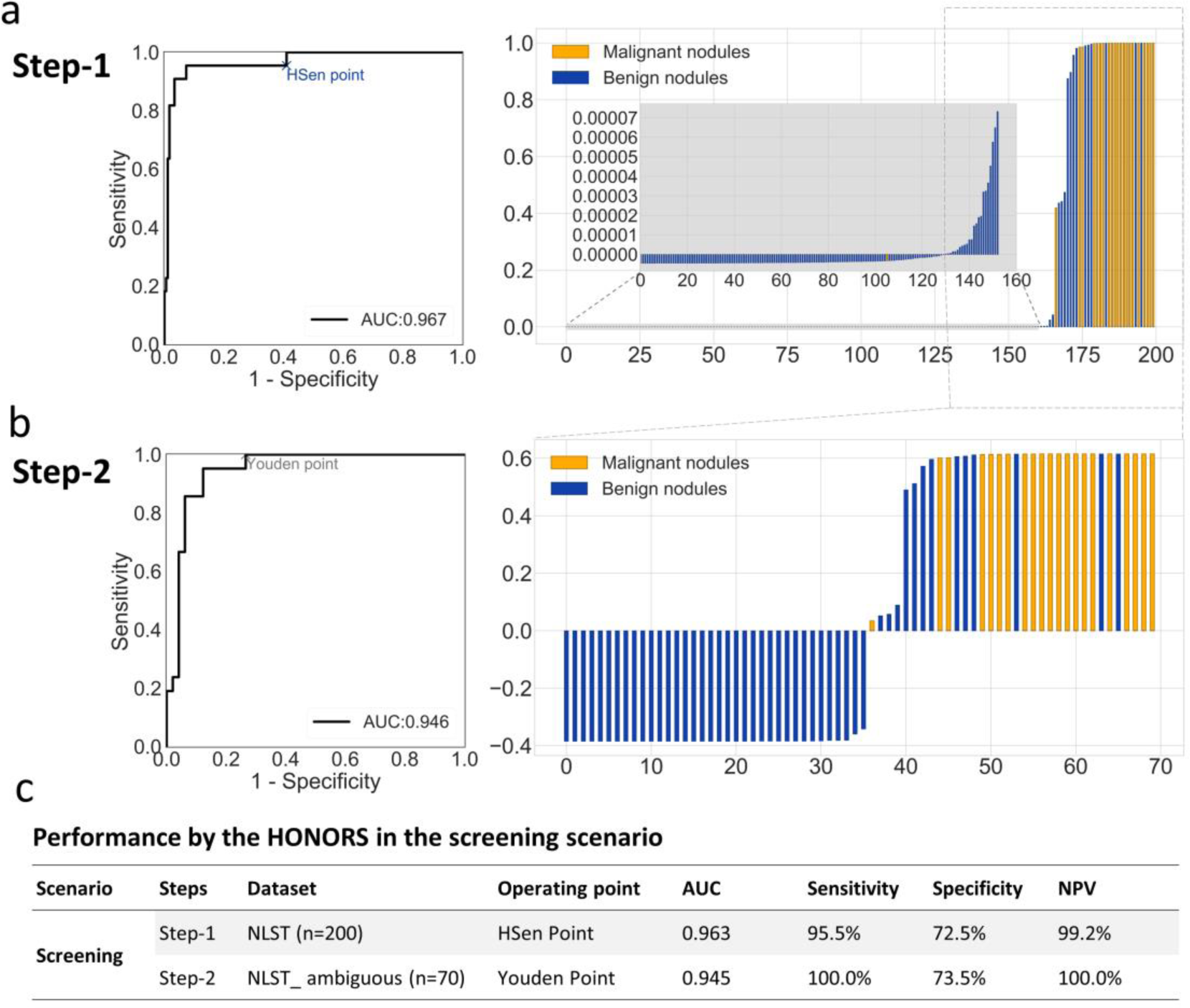
Performance of HONORS in screening scenario using NLST test set. a) In step 1, each line represents the relative malignant score (y-axis) of one nodule and x-axis represents the nodule index. Nodules with scores lower than 0.000005 were directly considered as benign nodules. The remaining ambiguous nodules will flow in to b) step 2 where each line represents the corresponding malignant score that extracted by Youden point of 0.384642. Nodules with scores less than zero were predicted as benign nodules and malignant ones for the rest. c) Corresponding statistics were used to evaluate the performance of HONORS in screening scenario, including AUC, sensitivity, specificity and NPV.

#### Evaluation of HONORS in diagnostic scenario

To incidentally detected nodules in routine diagnostic scenario (yellow path in Figure 1a), we took a HSpe point of FGP-NET and evaluated its performance on both the multi-center (n = 242) and JLH test set (n = 100), along with a reader comparison test based on JLH test set. FGP-NET achieved a high specificity of 94.5% (95% CI: 84.5-98.1%) and a sensitivity of 41.7% (95% CI: 34.7-48.9%) on multi-center set in step-1 with 81 nodules identified as lung cancer (Figure 5a, c). In step-2, a sensitivity of 93.6% (95% CI: 87.0-96.9%) and a specificity of 50.0% (95% CI: 36.3-63.1%) were achieved at the Youden point for the rest ambiguous nodules (Figure 5b, c). HONORS also exhibited great performance in JLH test set and outperformed the radiologists (Supplementary Figure S4, S5).

**Figure 5:**
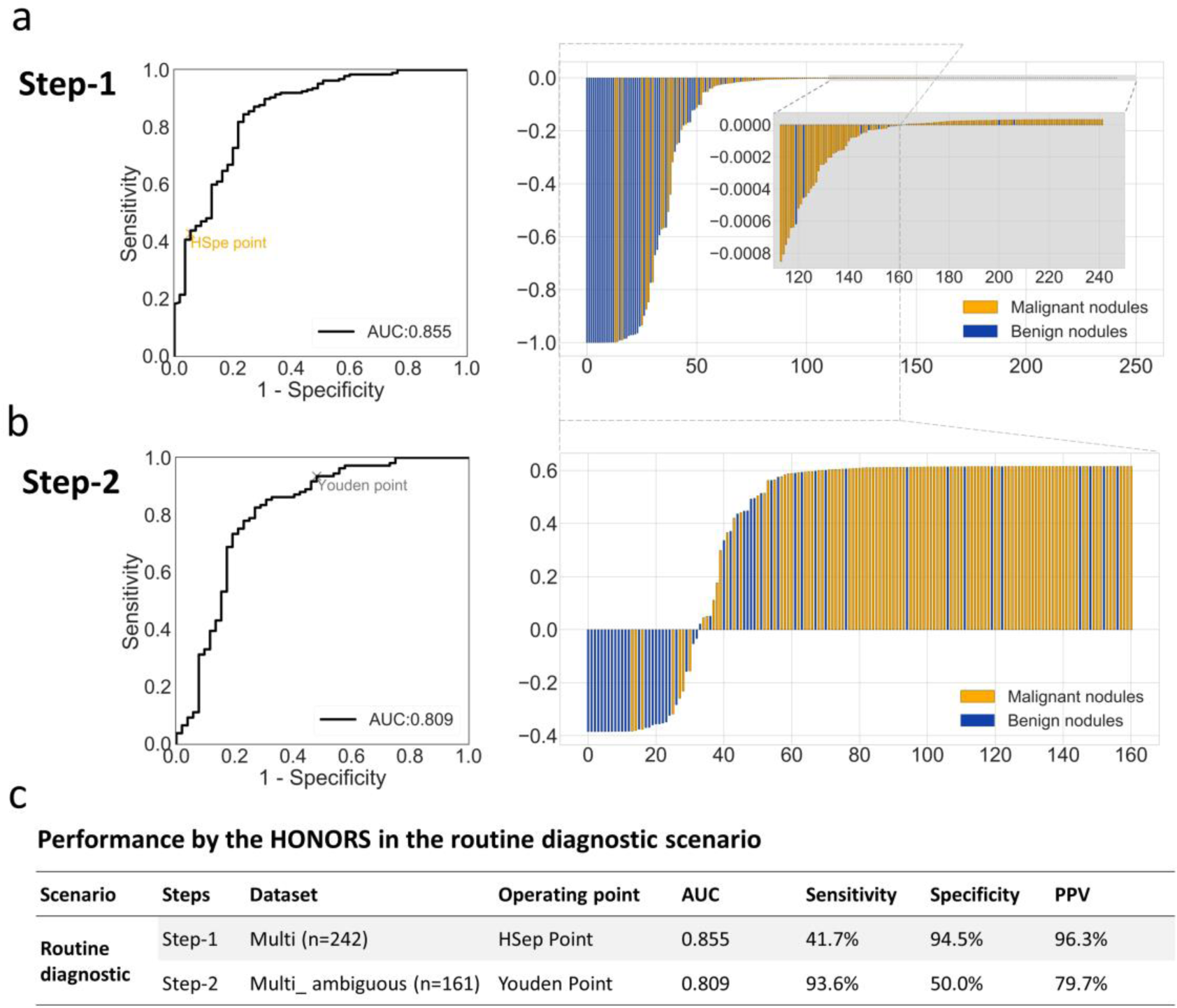
Performance of HONORS in routine diagnostic scenario using multi-center set. In step 1, each line represents the relative malignant score (y-axis) of one nodule in a) and x-axis represents the nodule index, and nodules with scores greater than 0.999965 were directly considered as malignant nodules. The remaining ambiguous nodules will flow into b) step 2 where each line represents the corresponding malignant score that extracted by Youden point of 0.384642. Nodules with scores greater than zero were predicted as malignant nodules and benign ones for the rest. c) Corresponding statistics were used to evaluate the performance of HONORS under routine diagnostic scenario, including AUC, sensitivity, specificity and PPV.

## Discussion

We developed and validated a DL algorithm—FGP-NET that is capable of stratifying pulmonary nodules with great performance and is comparable with a large group of radiologists. To further applied FGP-NET to different clinical settings, a novel two-step strategy—Hierarchical-Ordered Network-ORiented Strategy (HONORS) was proposed. It was able to stratify the nodules specifically based on different clinical scenarios (Hierarchical-Ordered) and the two-step workflow would become a possible stream for the application of DL in medical missions (ORiented). It may have potential to accelerate the process of pulmonary nodule diagnosis and to free doctors, nurses and other healthcare professionals to focus on providing real care for patients.

Our study was conceptually practical in clinics because FGP-NET and HONORS were designed to tailor both screening and routine diagnostic scenarios. Based on cutting edge technology of general image and fine-grained classification, FGP-NET was designed to effectively extract global features, multi-sized local features and complicated relationships among them, which significantly benefited the performance. Pyramid structure that we harnessed to support FGP-NET is a method for extracting multi-scale features and is quite suitable for feature extraction of pulmonary nodules due to its large size variation ^26^. Pyramid structure is typically deployed for detection and segmentation tasks but little is known about its performance in classification ^27^. To this end, we relied on pyramid structure and trained an automatic DL algorithm for pulmonary nodule risk prediction on single CT scan. High level of performance was achieved on internal test and it declined a little bit on external test probably due to the different nodule distribution and scanning, reconstruction parameters.

Radiomics approaches and DL have been applied universally for pulmonary nodule risk prediction in recent years ^28-30^. Most of these studies focused on the screening population. For example, Huang et al. developed a computer-aided diagnosis approach with a sensitivity of 95% and a specificity of 88% which outperformed three radiologists’ combined reading using 186 nodules from NLST dataset ^29^. Diego et al. ^14^ achieved the state-of-the-art performance (AUC = 0.944) for lung cancer risk prediction on 6,716 NLST cases. These researches suggested that DL-based model outperformed human experts in predicting lung cancer risk based on screened nodules. Our algorithm also exhibited great performance in the NLST dataset which represented screen nodules. Nevertheless, researches concerning incidentally detected nodules were limited and the performance of radiologists on them still remains unknown. Thus, we conducted a reader comparison study among a large group of 126 radiologists for incidentally detected nodules. In this study, FGP-NET exhibited the same accuracy rate and good agreement with the majority of the radiologists, which indicated that FGP-NET may have the potential to improve the individual level and diagnostic consistence of the radiologists.

Based on FGP-NET, we further proposed a two-step strategy—HONORS, which is promising to optimize clinical workflow and realize personalized precise treatment of pulmonary nodules. In the first step, the benign nodules or lung cancer can be accurately identified using HSen or HSpe point in screening or routine diagnostic scenarios, respectively. Such methodology of tuning high sensitivity or high specificity to match requirements for specific clinical settings has been presented in other areas like detection of diabetic retinopathy ^31^, screen for hyperkalemia ^32^, detection of moderate and large pneumothorax ^33^ to name but a few. In the screening scenario, approximately 96.4% of the nodules were benign at baseline screening ^34-36^, which brought excessive workload for physicians and unnecessary harm for patients from additional diagnostic procedures. Therefore, we set a HSen point by valuing sensitivity as 99% to stratify the benign nodules without any confirmation by physicians. FGP-NET made to stratify 75.2% of benign nodules with NPV at 99.2% in screening population. With such high NPV, FGP-NET could lead to accurate feedback for patients with examination of ‘benign nodules’ so that the precision could be improved. While in the routine diagnostic scenario, previous study indicated that 11.3% of patients underwent evaluation at more than one facility which would delay the treatment ^37^. Hence, a HSpe point by valuing specificity as 99% was set to stratify the lung cancer with high precision. In this context, FGP-NET stratified 41.7% of lung cancer with PPV at 96.3% in routine diagnostic population, which enabled examinations of ‘lung cancer’ to be moved ahead in the image interpretation workflow. Patients with lung cancer could receive timely diagnoses and treatment, even in primary hospitals where primary care providers are with low confidence and preference for referral ^38-40^. In the second step, further stratification of ambiguous nodules was performed to aid clinical decision making. Through HONORS, two management approaches of pulmonary nodules, that is, “Human-Free” and “Human-Machine Coupling” for clinical decision making can prosperously co-exist and may become the promising for the application of DL in real clinical settings.

However, this study has potential limitations. First, since all nodules from JLH dataset were scanned by Siemens, only images that were scanned by Siemens were selected from NLST to minimize the potential effect arose from different vendors; such practice biased the external validation due to the different involved vendors, resulting in a declined AUC. Future optimization should ensure the acquisition and reconstruction parameters of training data diversified to make the algorithm better in generalizing to an external dataset with different vendors. Second, the operating points were selected in validation set by valuing the sensitivity or specificity as 99%; however, the sensitivity or specificity failed to reach 99% at test set. Therefore, larger dataset was needed to train the algorithm and larger, prospective and multi-center validation is also necessary to examine the generalization of the algorithm towards a variety of population and clinical settings. Third, the radiologists were only informed of one CT scan while blind to the clinical information and prior CT scans in the contest, which was different from the real clinical routines. Therefore, further investigation and verification of our study are needed in the real-world setting.

Taken together, we proposed HONORS to lay groundwork toward application of the DL-based pulmonary nodule stratification algorithm in the screening and routine diagnostic scenarios. Future studies are warranted to prospectively assess the performance and generalizability of the algorithm at a variety of sites in real-world-use scenarios, and determine how the HONORS would impact diagnostic accuracy and clinical workflow.

## Data Availability

We used one dataset which is publicly accessible:
NLST: https://biometry.nci.nih.gov/cdas/learn/nlst/images/
Retrospective data used in this study from clinical hospitals cannot be released under the terms of our Institutional Review Board approval to protect patient confidentiality.

